# Encounters after Appointments Cancelled Due to COVID-19 in the Veterans Affairs Health Care System

**DOI:** 10.1101/2021.11.17.21266381

**Authors:** Linda Diem Tran, Liam Rose, Tracy Urech, Anita Vashi

## Abstract

This statistical brief examines subsequent encounters after a cancellation due to COVID-19 in the Veterans Affairs System. We find that he vast majority of VA patients that had appointments cancelled in mid-March to mid-April of 2020 had another encounter within 180 days. The most common next encounter was a virtual visit with a VA provider on the same day of the original appointment. We also find that patients that saw a provider through VA community care had a lower median time to next encounter.

## Introduction

The COVID-19 pandemic caused an unprecedented interruption in healthcare delivery. By mid-March 2020, nearly every healthcare system had postponed, delayed, or canceled many non-emergent services. VA was no different, experiencing substantial and lasting shifts in care delivery methods.^1^ There was widespread concern that patients may have been harmed through delays in needed care.^2,3^

This statistical brief presents data from the Veterans Health Administration (VA) on encounters after the initial wave of cancellations in March 2020. The report examines over 750,000 patients with appointments that were scheduled to occur with VA providers between March 11, 2020 and April 18, 2020 and were cancelled before the scheduled date and on or after March 11, 2020. It then shows where patients were seen next – if at all – within the 180 days after the date of the original appointment. Next encounters could be either inpatient or outpatient. A comparison group of patients – consisting of patients that did not have a appointment during this period, but had an encounter with VA within the previous year – is included to better contextualize the next encounter times. To calculate the median time to next encounter, these comparison patients were assigned a “cancellation date” of March 24, 2020, which was the date with the most actual cancellations.

## Findings

There were 750,190 patients with encounters cancelled during this period (Figure 1). Of these, the vast majority had an encounter within the next 180 days, with only 9 percent not seen in any capacity. Over 80 percent had an encounter with a VA provider. Other patients had a visit with VA community care, and only a small number died without another encounter within 180 days of the original appointment date.

**Figure 1.**
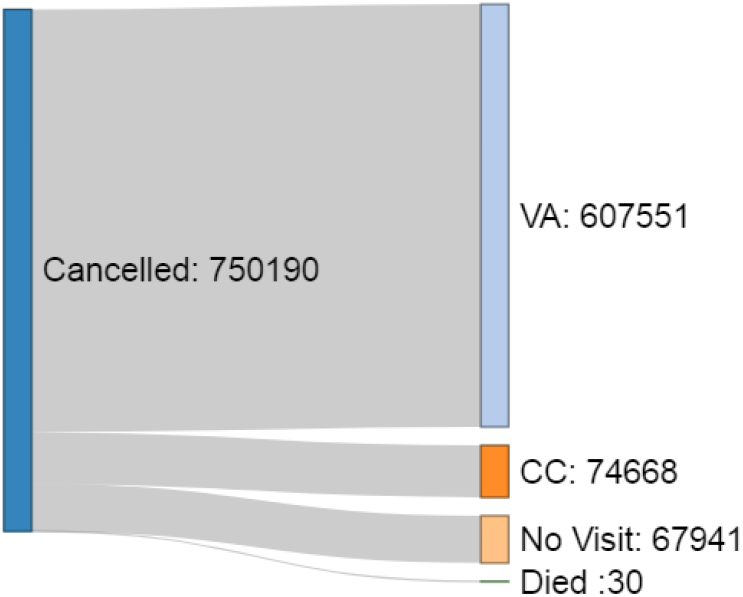
Diagram of Volume of Cancelled Appointments and Next Encounters *Notes: “Cancelled” refers to visits scheduled between March 11, 2020 and April 18, 2020 that were cancelled before the appointment and after March 11, 2020. “VA” refers to encounters with VA providers, “CC” refers to encounters with VA community care providers, and “No Visit” refers to no encounter within 180 days of the original appointment*.

Among patients that had a visit paid or provided by VA, the median time to their next encounter was 26 days (Figure 2). This number is somewhat higher for patients that had their encounter with a VA provider (23 days) than those that had their encounter through a community care provider (15 days).

**Figure 2.**
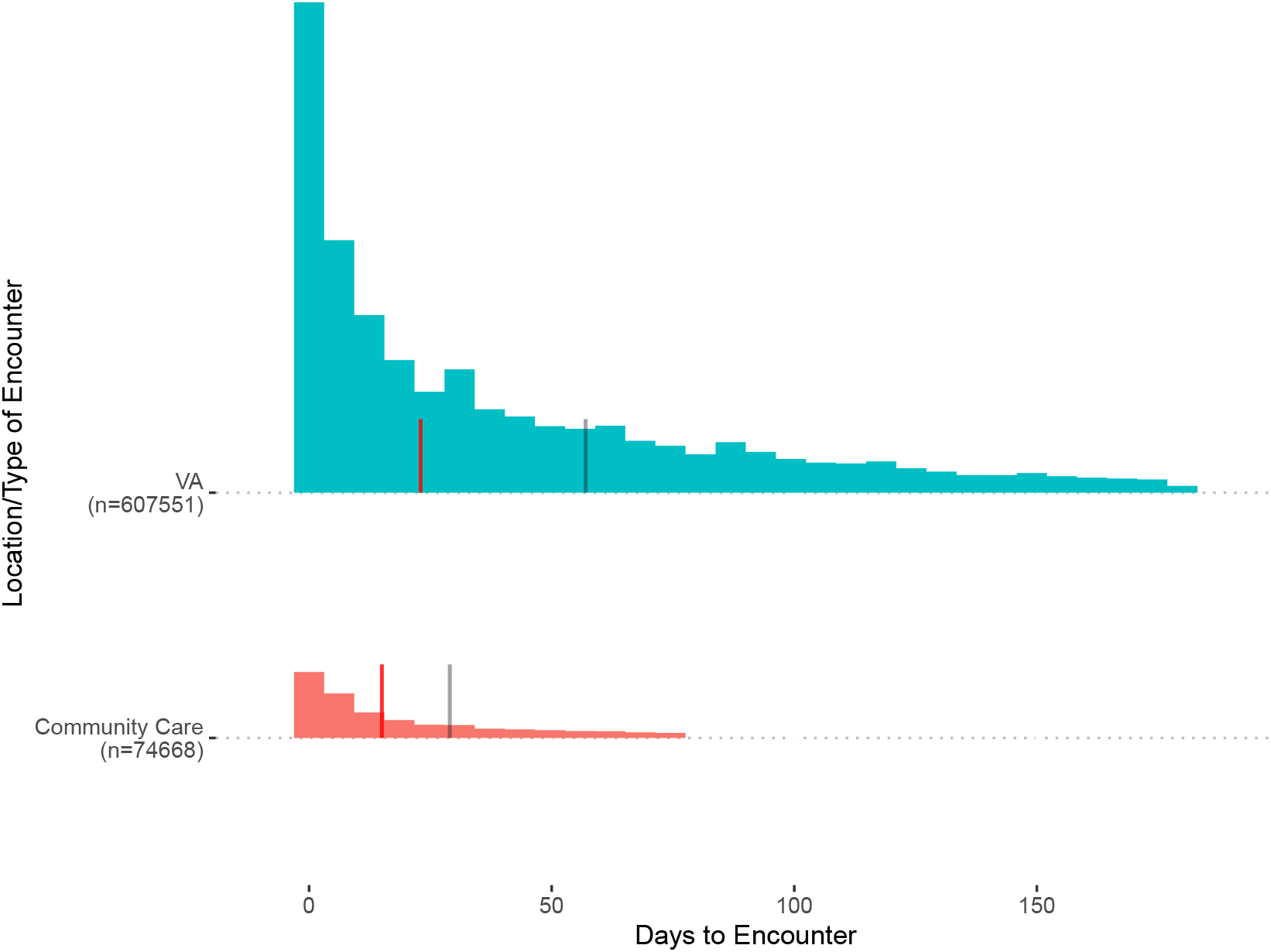
Time to Next Encounter from Date of Cancelled Appointment *Notes: “Days to Encounter” signifies the number of days between the patient’s originally scheduled appointment and their next encounter “VA” means the next encounter was with a VA provider, and “Community Care” means the encounter was with a community care provider. The red line signifies the median number of days to next encounter for each group. The gray line signifies the median number of days for the comparison group that did not have a scheduled appointment cancelled*.

Figure 3 further breaks down the type of next encounter for patients that had one within 180 days. The most common next encounter was a VA virtual visit on or near the date of the original appointment. Previous work has shown that VA was able to pivot to virtual care extremely quickly during this period.^4^

**Figure 3.**
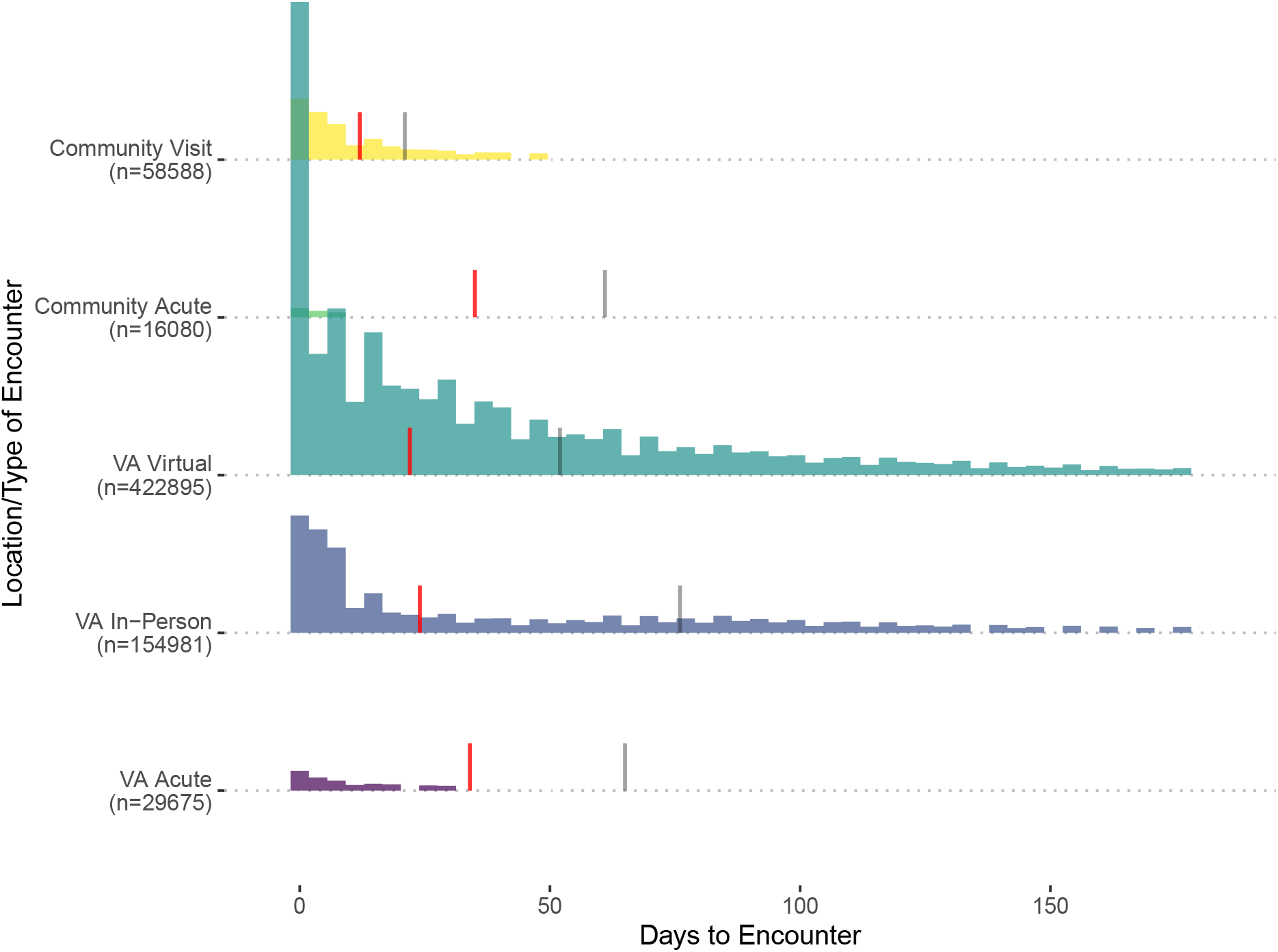
Diagram of Volume of Cancelled Appointments and Next Encounters *Notes: “Cancelled” refers to visits scheduled between March 11, 2020 and April 18, 2020 that were cancelled before the appointment and after March 11, 2020. “VA” refers to encounters with VA providers, “CC” refers to encounters with VA community care providers, and “No Visit” refers to no encounter within 180 days of the original appointment*.

**Table 1.**
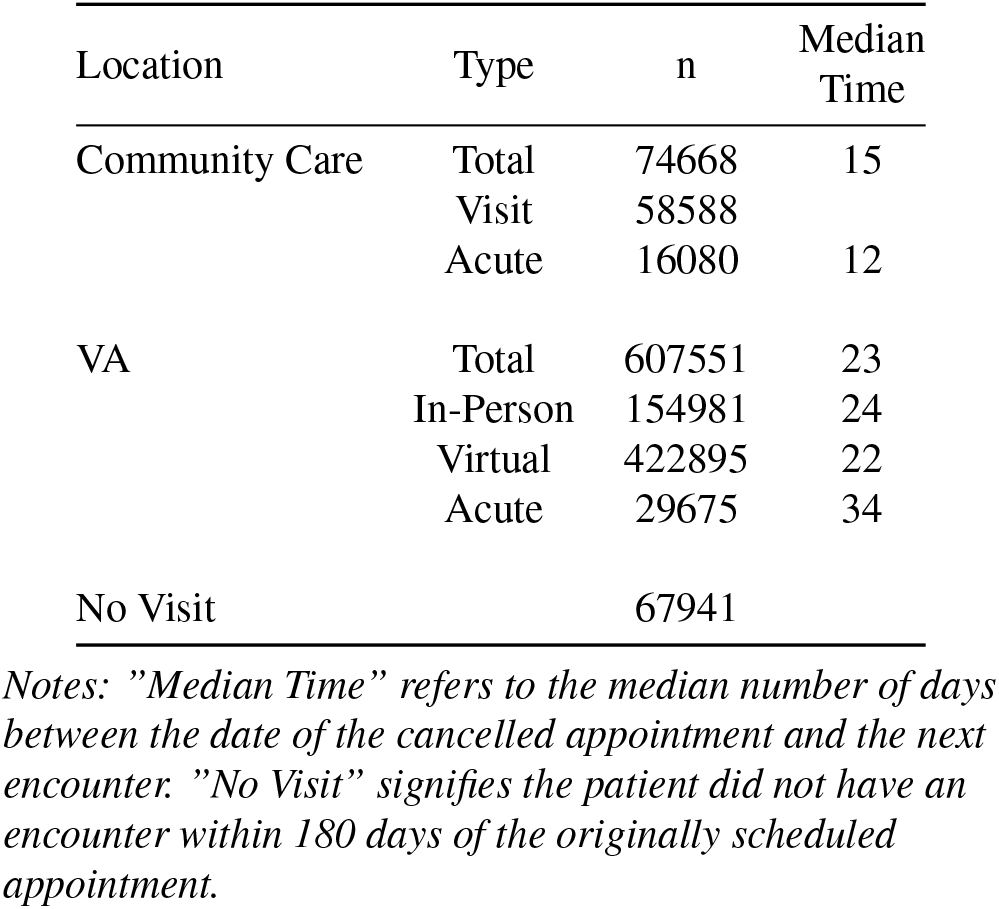
Volume and Median Time to Next Encounter by Location and Type of Encounter

The next most common next encounter was a VA in-person visit within two weeks of the originally scheduled appointment. Community visits were the next most common, and these visits had the shortest median time to from the scheduled appointment date. Only a relatively small fraction of patients had their next encounter in an acute setting, counted as any of urgent care, emergency department, or inpatient. This was true for both community care and VA provided care. In all cases, the median number of days to the next encounter was lower for patients with a cancelled appointment (red lines) than it was for patients without a cancelled appointment (gray lines).

## Limitations

These data do not take into account individuals that sought care outside VA and VA community care, such as care paid by Medicare or a private payer. Further, this report does not consider if the next encounter after initial appointment is for the same specialty or type of care. We included all cancellations during this period. Cancellation reasons included changes in medical condition and other non-health reasons, but inconsistencies in data entry during the early period of the pandemic prevented data from being sufficiently precise to differentiate from COVID-19 and other reasons for cancellation.^5^ Finally, we were unable to reliably differentiate community care in-person and virtual encounters.

## Data Availability

Data used in this analysis include personal health information and are not publicly available.

## Acknowledgments

The views expressed in this article are those of the authors and do not necessarily reflect the position or policy of the Health Economics Resource Center, the Department of Veterans Affairs, or the United States government.

